# Establishing the Automatic Identification of Clinical Trial Cohorts from Electronic Health Records by Matching Normalized Eligibility Criteria and Patient Clinical Characteristics

**DOI:** 10.1101/2024.02.28.24303396

**Authors:** K. Lee, Y. Mai, Z. Liu, K. Raja, T. Jun, M. Ma, T. Wang, L. Ai, E. Calay, W. Oh, E. Schadt, X. Wang

## Abstract

The use of electronic health records (EHRs) holds the potential to enhance clinical trial activities. However, the identification of eligible patients within EHRs presents considerable challenges. We aimed to develop a pipeline for phenotyping eligibility criteria, enabling the identification of patients from EHRs with clinical characteristics that match those criteria. We utilized clinical trial eligibility criteria and patient EHRs from the Mount Sinai Database. The criteria and EHR data were normalized using national standard terminologies and in-house databases, facilitating computability and queryability. The pipeline employed rule-based pattern recognition and manual annotation. Our pipeline normalized 367 out of 640 unique eligibility criteria attributes, covering various medical conditions including non-small cell lung cancer, small cell lung cancer, prostate cancer, breast cancer, multiple myeloma, ulcerative colitis, Crohn’s disease, non-alcoholic steatohepatitis, and sickle cell anemia. 174 were encoded with standard terminologies and 193 were normalized using the in-house reference tables. The agreement between automated and manual normalization was high (Cohen’s Kappa = 0.82), and patient matching demonstrated a 0.94 F1 score. Our system has proven effective on EHRs from multiple institutions, showing broad applicability and promising improved clinical trial processes, leading to better patient selection, and enhanced clinical research outcomes.

## Introduction

Patient recruitment and retention pose significant challenges in the clinical trial domain ^1^, with poor accrual rates often leading to trial failures. Challenges range from disease-specific issues, like the rarity of conditions ^2,3^, to systemic issues such as market competition, lack of knowledge, uncertainties of patients about being a study subject ^3,4^, and rigid protocols that disqualify many from participation ^5^. Moreover, the traditional manual medical record review for patient selection is excessively burdensome and often unviable for assessing large cohorts. A potential solution to these challenges is the strategic application of technology for the expedited pre-screening of eligible patients through Electronic Health Records (EHRs), an approach that has demonstrated notable improvements in the efficiency and precision of identifying appropriate trial cohorts ^6–10^.

Electronic clinical phenotyping, which extracts clinical features and patient characteristics from large datasets plays a pivotal role in precision and population-based medicine ^11–13^. It serves as an instrumental tool for the selection of cohorts for clinical predictive modeling, identification of clinical trial cohorts, and evaluation of healthcare quality ^14^. Converting clinical trial eligibility criteria (EC) into computer-interpretable formats has facilitated the identification of clinical phenotypes necessary for various applications, including cohort selection ^4,5,8,15^. Despite the potential benefits, automated clinical phenotyping from EHR data faces several challenges ^16^. Earlier efforts have focused on parsing clinical trial EC into formats that computers can interpret to support trial protocol design ^17^, automated cohort selection, and collaborative clinical research ^18–24^. Computer languages such as Arden Syntax ^25^, Guideline Expression Language Object-Oriented (GELLO) ^26^, ECLECTIC ^27^, and Clinical Trail Markup Language ^28^, have been developed to represent EC in a way that machines can process. Template-based approaches like Eligibility Rule Grammar and Ontology (ERGO) ^29^ and Eligibility Criteria Extraction and Representation (EliXR) ^30^, transform EC into computable representations. These computable representations can be applied in various database query languages such as SPARQL^31^, Web Ontology Language (OWL) Description Logics (DL) ^32,33^, and SQL to automate clinical phenotyping ^17^. Tools like Criteria2SQL convert EC directly into SQL queries ^17^ and certain approaches organize structured EC according to the Observational Medical Outcomes Partnership (OMOP) Common Data Model (CDM) ^34,35^. Despite these advancements, these methodologies encounter limitations due to the reliance on natural language-based syntax, which may introduce errors resulting from abbreviations and typographical errors in clinical terms. Furthermore, the accessibility of the syntax presents challenges.

In this study, we addressed these limitations by building an advanced intermediate representation of normalized and standardized clinical concepts that enhance implementation via SQL queries. Our clinical phenotyping pipeline comprises three components. Firstly, we developed a rule-based knowledge engineering component to annotate the EC attributes into a computable and customizable granularity from EHRs. Secondly, we normalized the heterogeneity of clinical expressions in the annotated EC attributes and EHRs to predefined medical concepts from standard terminologies and four in-house knowledge bases (procedures, medications, biomarkers, and diagnosis modifiers). Thirdly, we constructed a knowledge base of computable criteria attributes to match patients to clinical trials. This knowledge base can support a range of purposes, including cohort selection and trial protocol design.

## Materials and methods

### Data Sources

Data were sourced from ClinicalTrials.gov (https://clinicaltrials.gov/) and EHR from Sema4 data warehouse, which includes the Mount Sinai Data Warehouse (MSDW) and VieCure, a next-generation clinical decision support platform (https://www.viecure.com/). The EHR data encompassed comprehensive patient information including patient demographics, vital signs, medical histories, diagnoses, medications, lab test results, immunization dates, allergies, and radiology images. The study is covered under IRB-17-01245 approved by the Program for the Protection of Human Subjects at the Mount Sinai School of Medicine.

We utilized the EC attributes extracted from a total of 3,475 clinical trials. The trials included 3,281 previously analyzed trials, covering non-small cell lung cancer, prostate cancer, breast cancer, multiple myeloma, ulcerative colitis, and Crohn’s disease, leveraging a deep-learning-based NLP technique ^15^. An additional 194 trials recruiting small cell lung cancer, non-alcoholic steatohepatitis, and sickle cell anemia were analyzed before clinical phenotyping. All extracted EC attributes as well as patients’ clinical characteristics retrieved from EHR were categorized into ten clinical domains: condition, procedure, lab test, therapy, biomarker, diagnosis modifier, observation, line of therapy, vital signs, and demographic. EC and EHR data were normalized to establish the mapping and then saved in a knowledge base for further analysis and reference.

### Clinical Phenotyping Pipeline

Our pipeline comprises three key components: 1) Rule-based knowledge engineering, 2) Normalization of EC attributes and clinical characteristics, and 3) Clinical phenotyping knowledge base.

#### Rule-based knowledge engineering

Therapy-related data from EHRs and trial EC were classified into five categories: (i) treatment (e.g. neoadjuvant therapy), (ii) regimen (e.g.TCH), (iii) modality (e.g. chemotherapy), (iv) mechanism of action (MOA) (e.g. EGFR inhibitor) and (v) medication (e.g. carboplatin). Standard resources and in-house knowledge bases were utilized for this purpose. Treatment, regimen, modality, and MOA were then mapped to specific medications using dedicated resources such as Cancer Alteration Viewer (CAV) and disease treatment guidelines (see Supplemental Table 1 for details). For example, MOA, anti-androgen for prostate cancer is mapped to several medications including bicalutamide, flutamide, nilutamide, apalutamide, darolutamide, enzalutamide, and abiraterone. For lab tests, biomarkers, and observations, we annotated attribute names and values. The groups were added to the EC attributes before saving them to the knowledge base (Fig. 1). Certain biomarkers within the EC attributes (e.g., HER2 R678Q) do not need further annotation while certain biomarkers (e.g., EGFR mutations sensitized to tyrosine kinase inhibitor) need annotation before mapping. We annotated such biomarkers with all possible mentions from the literature and examples mentioned in EC (e.g., L858R in exon 21, L861Q in exon 21, in-frame deletions in exon 19) to ensure comprehensive coverage. Medication classes (e.g. LHRH agonist) were annotated with corresponding medication (e.g., goserelin, leuprolide).

**Fig. 1.**
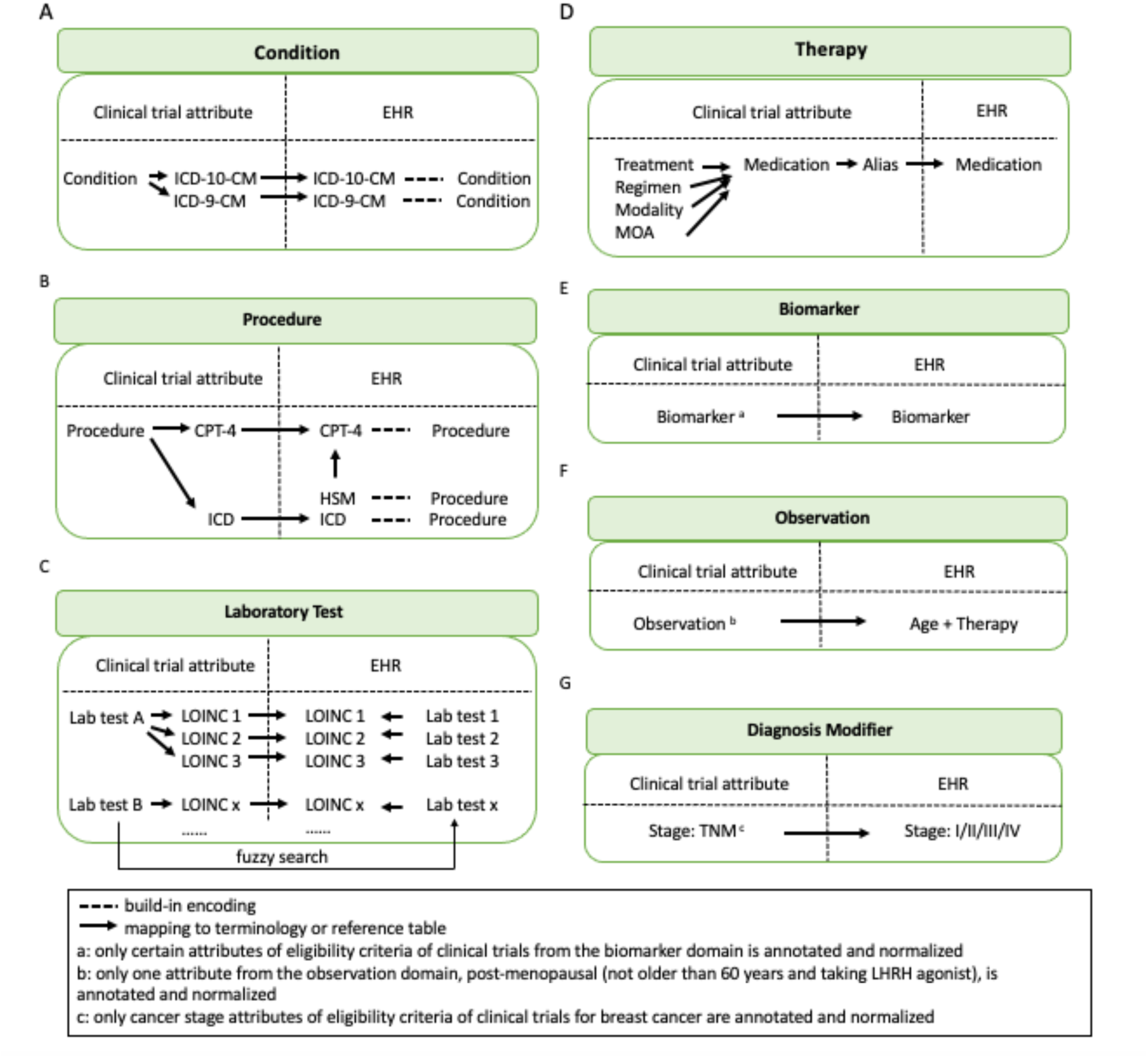
Clinical trial eligibility criteria phenotyping. (A) Eligibility criteria attributes from the condition domain are annotated, normalized, and mapped to clinical characteristics in EHR. (B) Eligibility criteria attributes from the procedure domain and clinical characteristics in EHR are annotated and normalized. Annotated and normalized attributes of eligibility criteria of clinical trials are mapped to normalized clinical characteristics in EHR. (C) Eligibility criteria attributes from the lab test domain are annotated and mapped to clinical characteristics in EHR from the lab test domain. Clinical characteristics in EHR from the lab test domain are annotated and normalized. Attributes of eligibility criteria of clinical trials are normalized through mapping to annotated and normalized clinical characteristics in EHR. (D) Eligibility criteria attributes from the therapy domain are annotated, normalized, and mapped to clinical characteristics in EHR. (E) Certain eligibility criteria attributes from the biomarker domain are annotated, normalized, and mapped to clinical characteristics in EHR. (F) Certain eligibility criteria attributes from the demographic domain are annotated, normalized, and mapped to clinical characteristics in EHR. (G) Certain eligibility criteria attributes from the diagnosis modifier domain are annotated, normalized, and mapped to clinical characteristics in EHR.

#### Normalization of EC attributes and clinical characteristics

We normalized EC attributes and clinical characteristics within the seven clinical domains, condition, procedure, lab test, therapy, biomarker, observation, and diagnosis modifier using standard resources such as International Classification of Diseases (ICD) 9^th^ and 10^th^ revisions, Current Procedural Terminology (CPT) 4^th^ edition, Logical Observation Identifiers Names and Codes (LOINC), and in-house knowledge bases (Fig. 1). The procedures mentioned in EHR are from two sources, the post-surgery documentation system from the EPIC database and Horizon Surgical Manager (HSM). The procedures from HSM are encoded with HSM code. We created an in-house knowledge base to map HSM code to CPT. The procedures from EPIC are either encoded with CPT, ICD-9, or ICD-10, or not encoded. We mapped the procedures without encoding in EPIC to CPT using the bioportal site (https://bioportal.bioontology.org/ontologies/CPT).

EC attributes and clinical characteristics from EHR within the lab test domain were normalized using LOINC codes (https://loinc.org/) (Fig. 1C). The system (e.g., serum), quantity (e.g., molar), time (e.g., mol/24h), type of scale (e.g., quantitative), and type of method (e.g., immunoassay) from a lab test were used for mapping it to the best LOINC code. For each lab test from the EC, we performed a fuzzy search to retrieve a list of related lab tests from EHR and normalized them to LOINC codes. The lab tests (e.g., C-reactive protein) without system, quantity, time, type of scale, and type of method may map to multiple lab tests in EHR (e.g., C reactive protein, C reactive protein HS). Normalization of each lab test from EHR may map to multiple LOINC codes (e.g., LOINC codes, 1988-5, 14634-0, 11039-5, and 76485-2 for c reactive protein; LOINC codes, 30522-7 35648-5, 76486-0 and 59182-6 for c reactive protein HS). To simplify the mapping, we defined a set of rules to map each lab test in EHR to one LOINC code (e.g., 1988-5 for c reactive protein and 30522-7 for c reactive protein HS):

Rule 1. Mapping the most popular lab test in the LOINC dictionary to the lab test in EHR, when the popularity rank is available in the LOINC dictionary.

Rule 2. Mapping the lab test for serum and/or plasma samples in the LOINC dictionary to the lab test in EHR, when the popularity rank is not available in the LOINC dictionary.

Rule 3. When one-to-one mapping is not possible with Rule 1 and Rule 2, the test unit is applied to achieve the mapping.

Rule 4. When one-to-one mapping is not possible with Rule 1, Rule 2, and Rule 3, the unit gram is preferred than molar for mapping.

Rule 5. When one-to-one mapping is not possible with Rule 1, Rule 2, Rule 3, and Rule 4, the lab test without information about method is preferred for mapping.

A medication within the therapy domain can be mentioned with different synonyms across multiple EHR records. We normalized the medications by retrieving all the synonyms (i.e., generic name, brand name, and abbreviation) from Unified Medical Language System (UMLS) Metathesaurus^35^ and RxNorm (https://mor.nlm.nih.gov/RxNav/). We observed that certain clinical characteristics from EHR within the diagnosis modifier domain were missing important information. For example, the breast cancer mentioned in EHR data from MSDW contains only clinical stages like I, II, III, or IV, not TNM stages. We normalized such clinical characteristics with the missing information (e.g., T1N0M0 = stage I) based on the National Comprehensive Cancer Network (NCCN) guidelines. Additionally, the “early stage, advanced stage, and metastasis” stages in EC attributes were normalized to the clinical stages. Normalization of example EC attributes and clinical characteristics is shown in Table 1.

To address the challenge of exact matching between EC attributes and clinical characteristics, we implemented two rules:

Rule A: We mapped EC attributes to clinical characteristics at a higher or lower level within EHR or standard terminologies. For instance, the attribute “interstitial lung disease” could be mapped to more specific concepts in ICD-10, such as acute respiratory distress syndrome, pulmonary edema, pulmonary eosinophilia, or other interstitial pulmonary diseases.

Rule B: We accounted for cases where an EC attribute is part of a clinical characteristic, or standard terminologies include additional details. For example, the attribute “colectomy” could correspond to a clinical characteristic like “colectomy/total/ostomy” or a standard terminology entry like “Colectomy, total; abdominal, without proctectomy; with ileostomy or ileoproctostomy.”

**Table 1.**
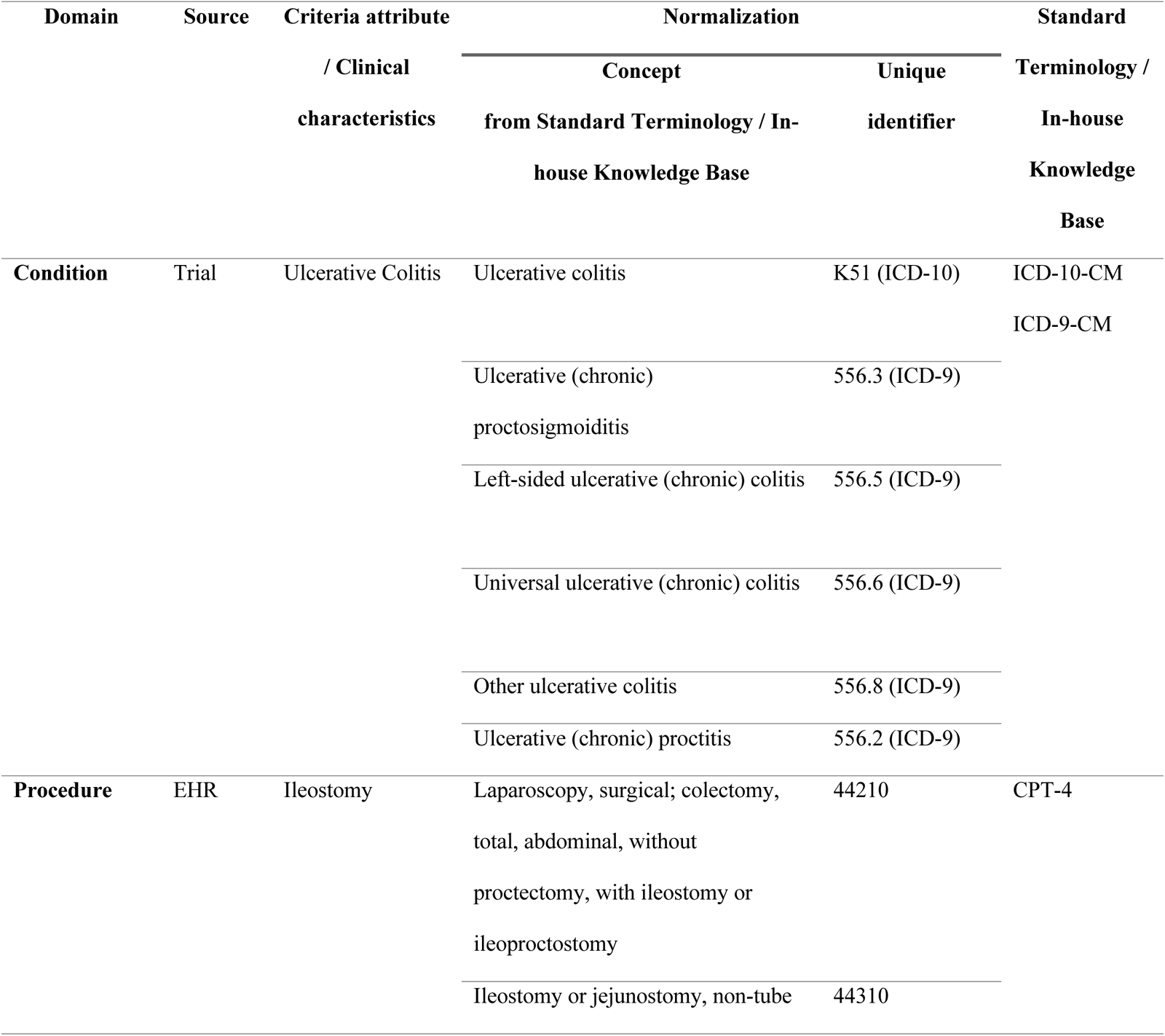
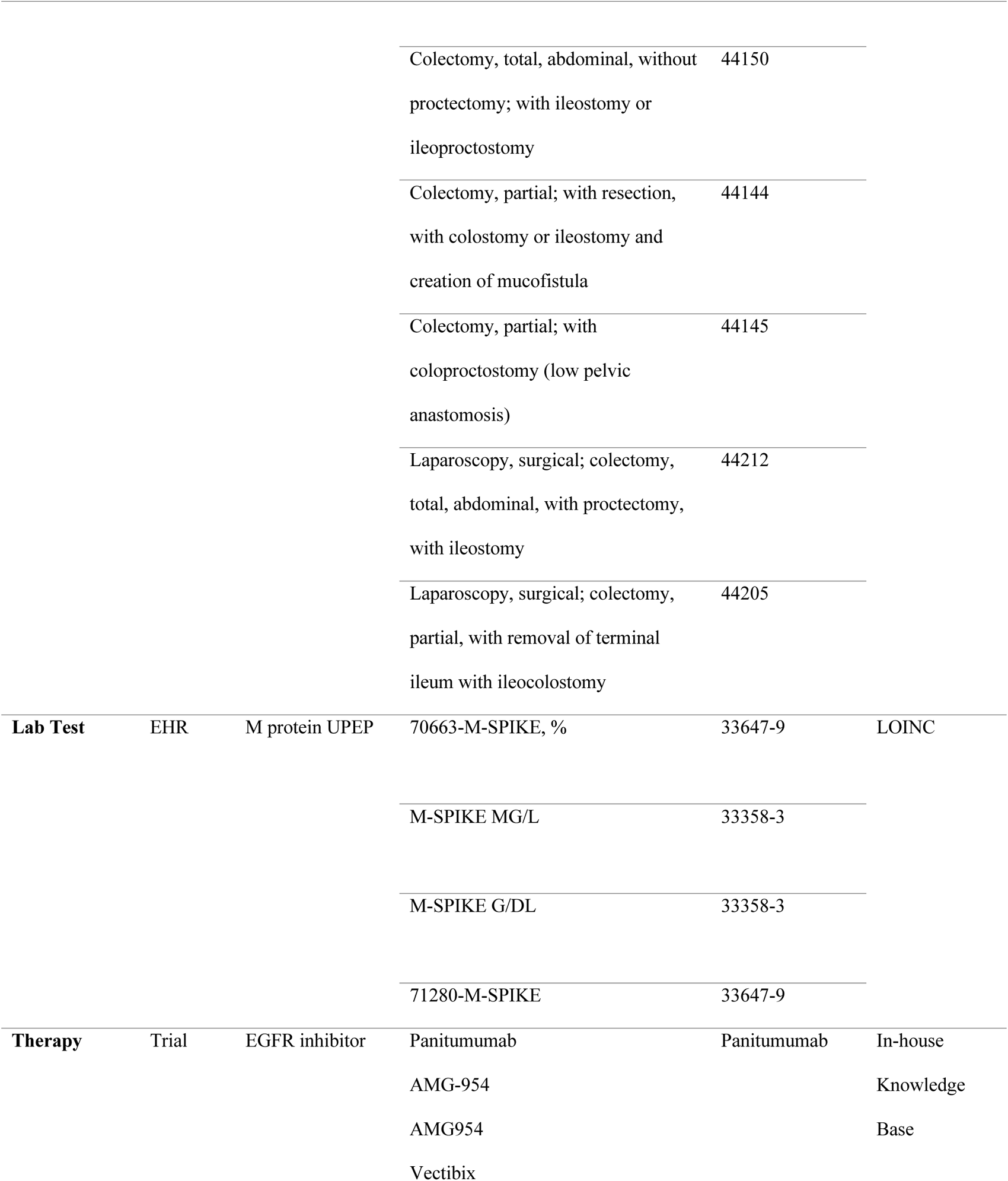
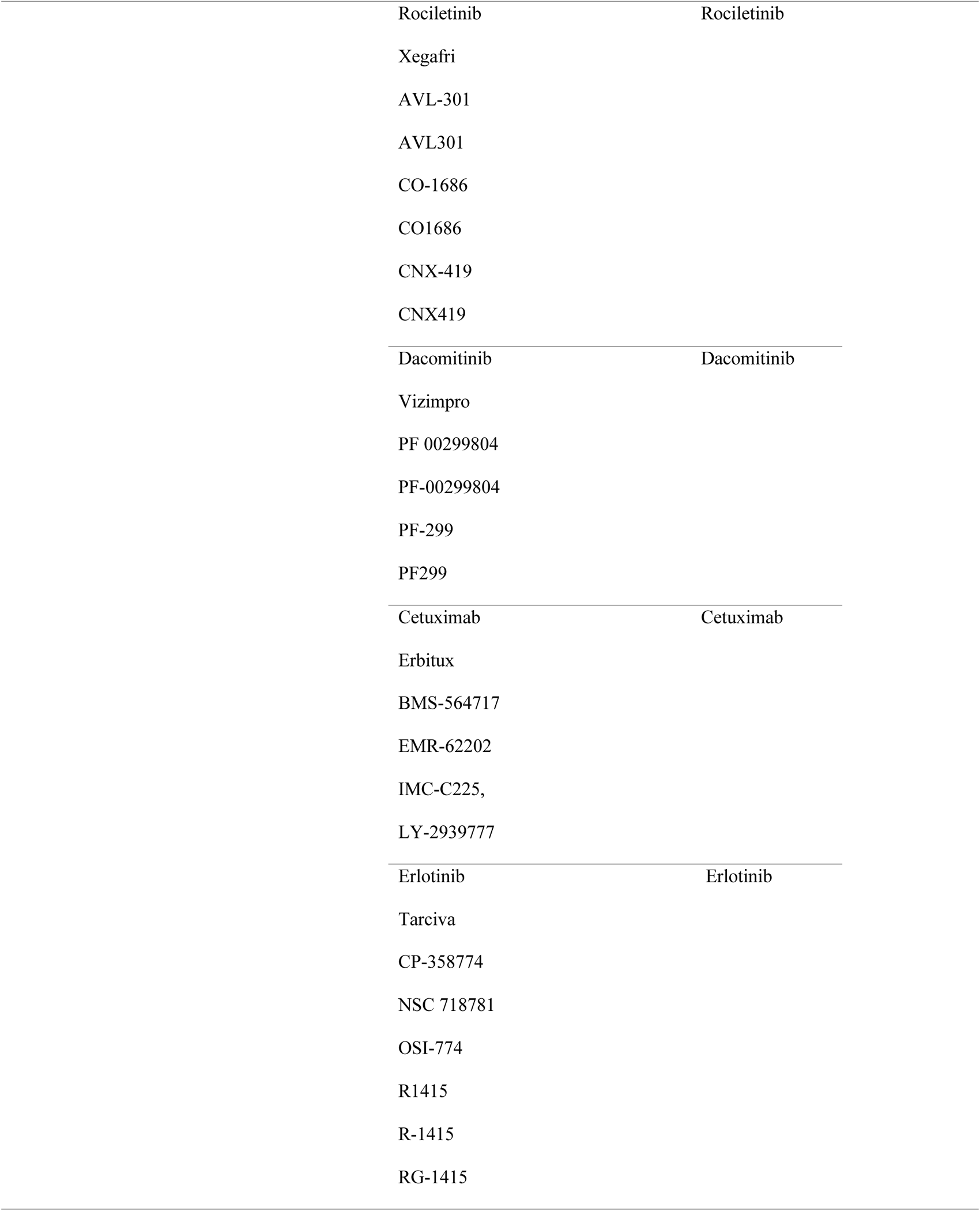
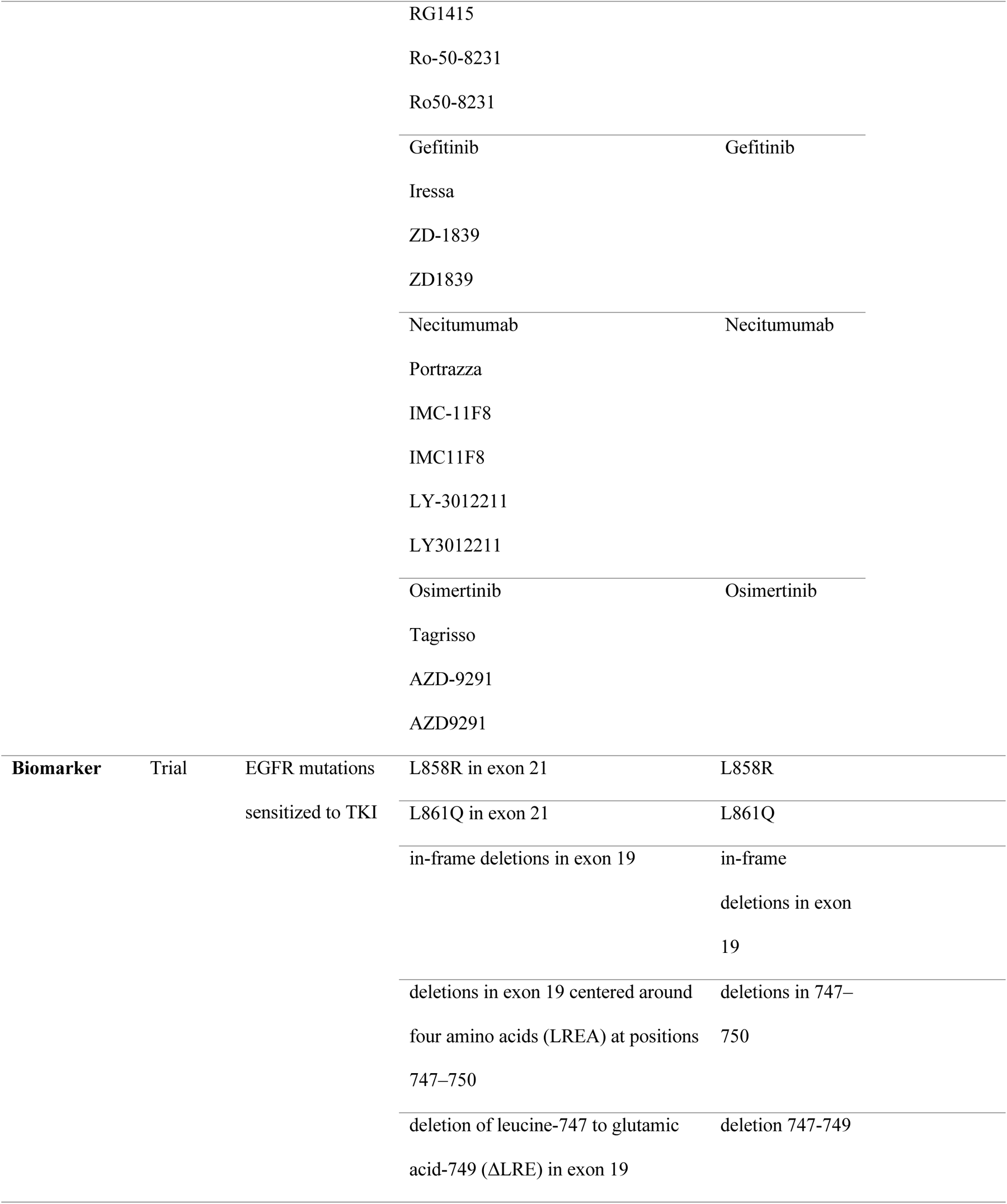
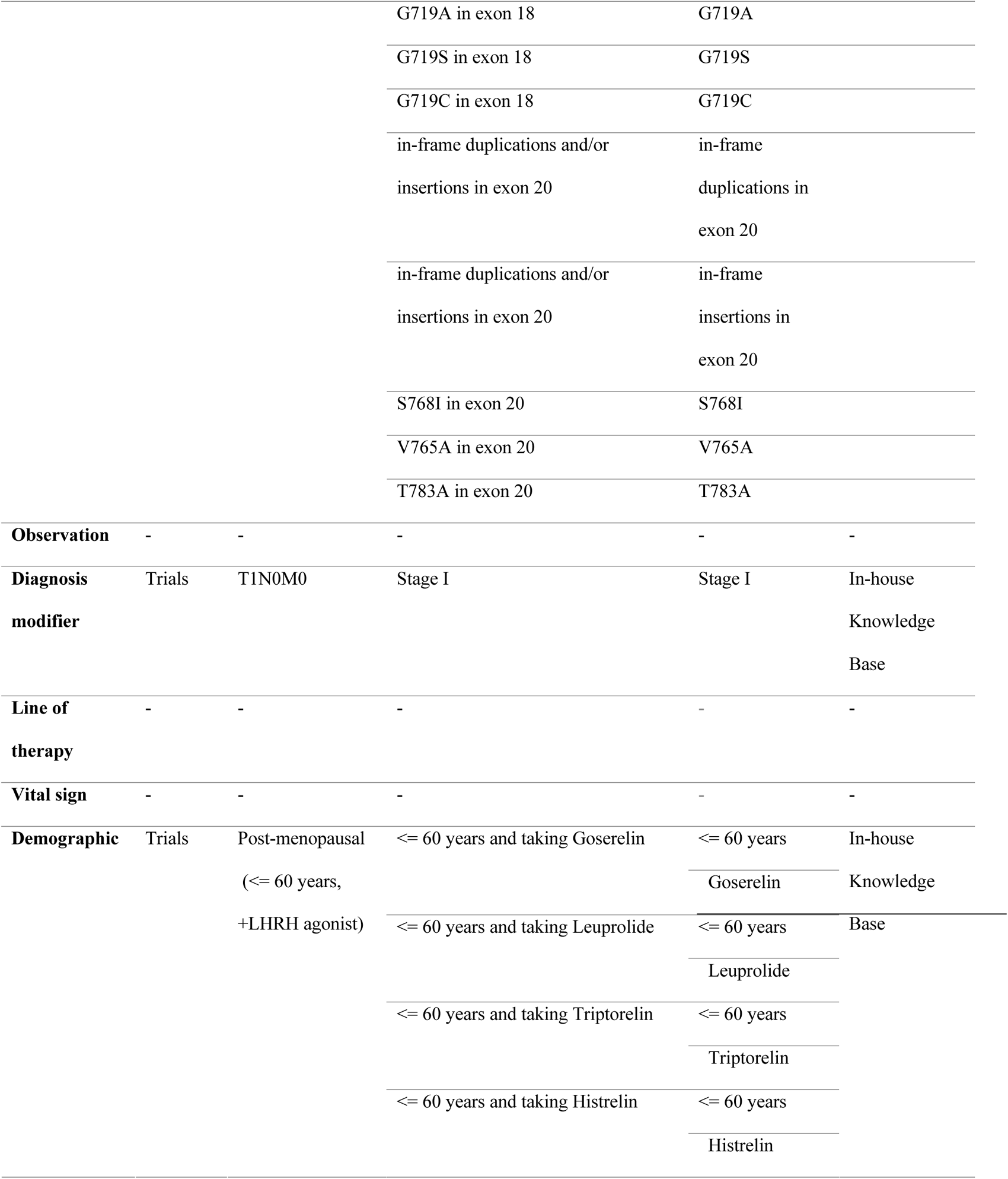
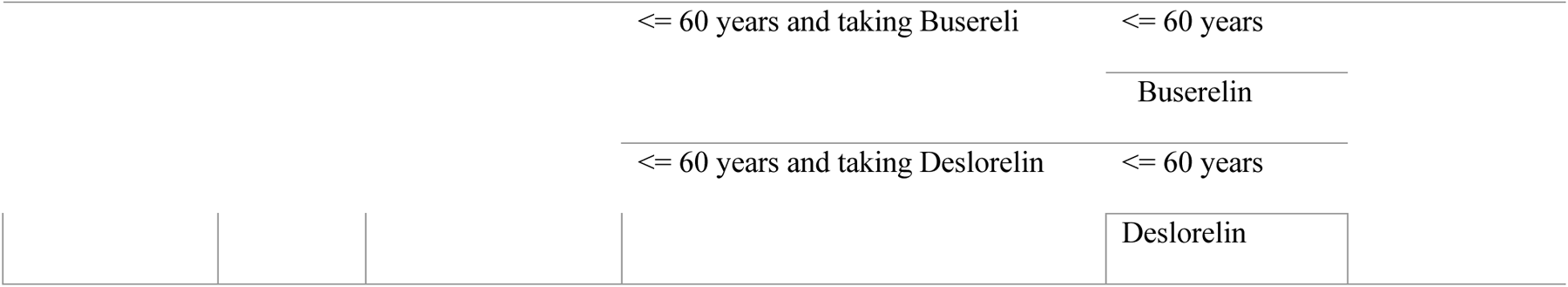
Example annotation and/or normalization of CT attributes and clinical characteristics.

### Clinical phenotyping knowledge base

The annotated and normalized EC attributes were indexed and stored in a Redshift database. The normalized clinical characteristics from EHR were also stored in the Redshift database as reference tables. The indexed EC attributes and reference tables together form the knowledge base for clinical phenotyping.

### Quality Control and Evaluation

We conducted quality assurance on a subset of annotated and normalized EC attributes and EHR clinical characteristics in the domains of condition, procedure, lab test, and therapy. For the condition domain, we ensured the appropriateness of mapped ICD-9 or ICD-10 codes for EC attributes, making necessary reassignments where needed. In procedure and lab test domains, we verified the correctness of mappings and LOINC codes between EC attributes and EHR clinical characteristics. Corrections were made where needed, following defined rules. Regarding the therapy domain, we reviewed the medication list for completeness and accuracy. Adjustments were made, such as removing medications like Lapatinib that have dual roles as EGFR and HER2 inhibitors.

In biomarkers, observations, and diagnosis modifiers domains, we reviewed the annotation completeness and correctness for each attribute, making updates based on careful examination. For instance, additional single-point substitutions in EGFR were added to the mutation list for EGFR mutations sensitized to tyrosine kinase in non-small cell lung cancer. Through the implementation of these rules and thorough reviews, we ensured the quality and accuracy of the annotated and normalized EC attributes and EHR clinical characteristics, enhancing the reliability of the data for further analysis and research.

Our evaluation involved two methods. First, a subset of the annotation from the Redshift database was assessed by two curators (YM and KL), with inter-rater agreement gauged by Cohen’s Kappa coefficient^31^. Second, we compared a random sample of expert-annotated EC attributes against a gold standard derived from EHR data (e.g., patient age at the time of phenotyping, the diagnosed conditions before the phenotyping was performed), measuring performance with precision, recall, and F1-score metrics.

## Results

### Annotated and normalized attributes

We extracted 640 unique attributes with values from 3,475 clinical trials (the whole list is provided in Supplemental Table 2) and grouped them under 10 clinical domains (Table 2). 367 out of 640 attributes (57.34%), belonging to seven clinical domains, condition, procedure, lab test, therapy, biomarker, observation, and diagnosis modifier, were annotated and normalized before storing in the Redshift database (See Supplemental Table 3 for details). Among the 363 annotated and normalized attributes, 174 attributes (47.41%) were normalized using the standard terminology, and 193 attributes (52.59%) were normalized using the concepts from the reference tables (Supplemental Table 2). While 72 attributes under lab tests were normalized using the standard terminology alone, two attributes under biomarker and one attribute under observation were normalized using the reference tables only. Normalization of attributes under therapy and diagnosis modifiers was mainly achieved with the reference tables. In the therapy domain, three attributes were normalized using standard terminology, and 163 attributes were normalized using reference tables. In the diagnosis modifier domain, seven attributes were normalized using standard terminology, and 18 attributes were normalized using reference tables. Our results show that EHR includes several attributes that are not in standard terminologies such as CPT, ICD-9-CM, and ICD-10-CM. The gap between EHR and the standard terminologies was filled with our reference tables. We did not annotate or normalize 273 attributes (42.66%) because 133 attributes (48.72%) did not require annotation or normalization (e.g., age), 140 attributes (51.28%) were difficult to achieve mapping to EHR (e.g., disease status “in remission/respond” and “unresolved toxicity from the prior treatment”).

**Table 2.**
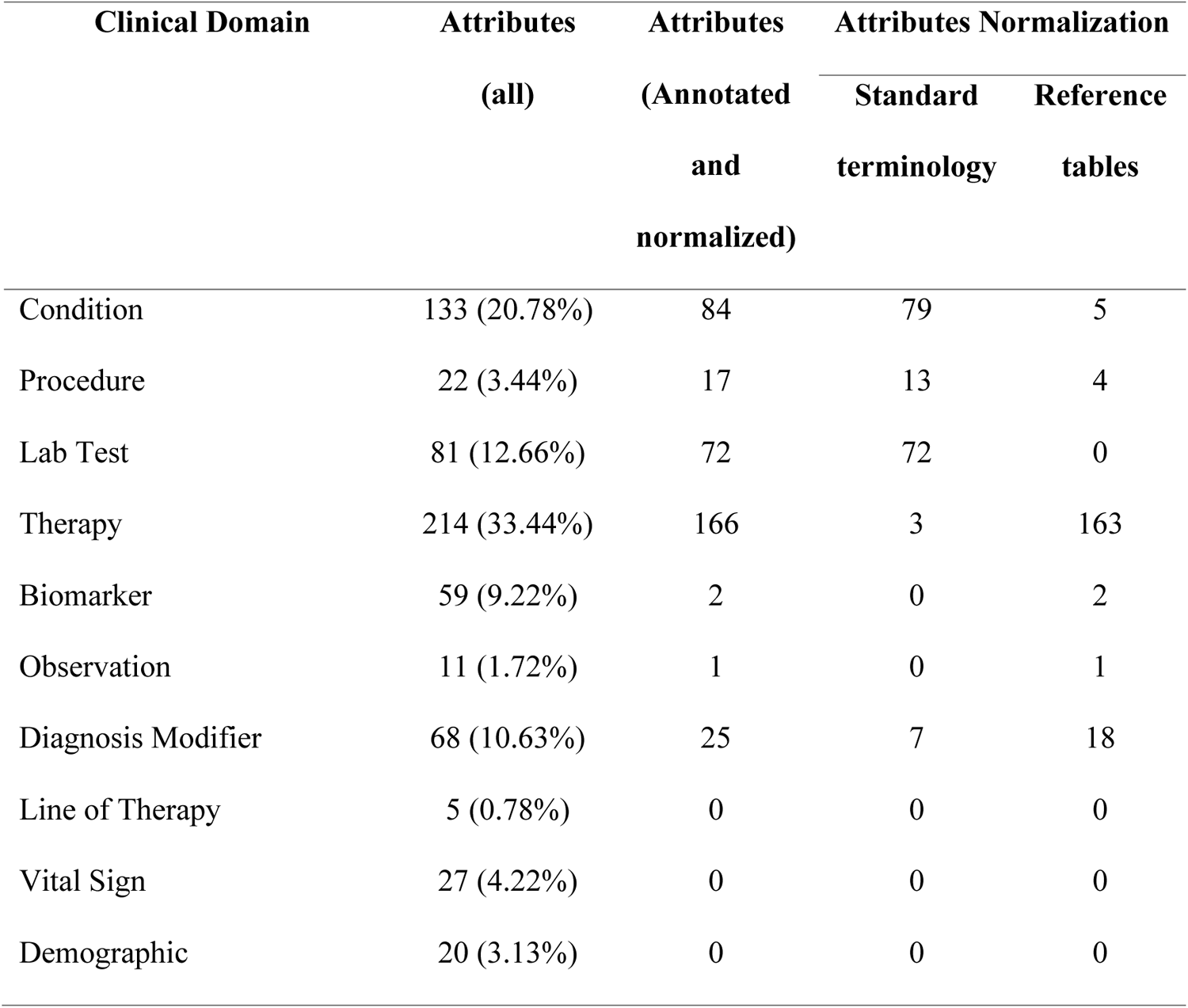
Annotated and normalized attributes of the eligibility criteria of clinical trials and the clinical characteristics of EHR.

### Attributes Distribution

The majority of the annotated and normalized EC attributes are from three domains: condition, lab test, and therapy. These attributes are dominantly found in clinical trials for non-small cell lung cancer, prostate cancer, and breast cancer (Fig. 2). Conversely, the unannotated and unnormalized attributes belong to seven groups: demographic, disease index, line of therapy, neoadjuvant treatment, radiotherapy, vital, and other (See Supplemental Table 4 for details). The annotated and normalized attributes of the EC of clinical trial belong to 28 attribute groups (Fig. 3A). Among, the four groups, test, targeted therapy, hormone therapy, and medication, were frequently mentioned in the EC of the clinical trials (i.e., 58.31% of all annotated clinical trial attributes).

**Fig. 2.**
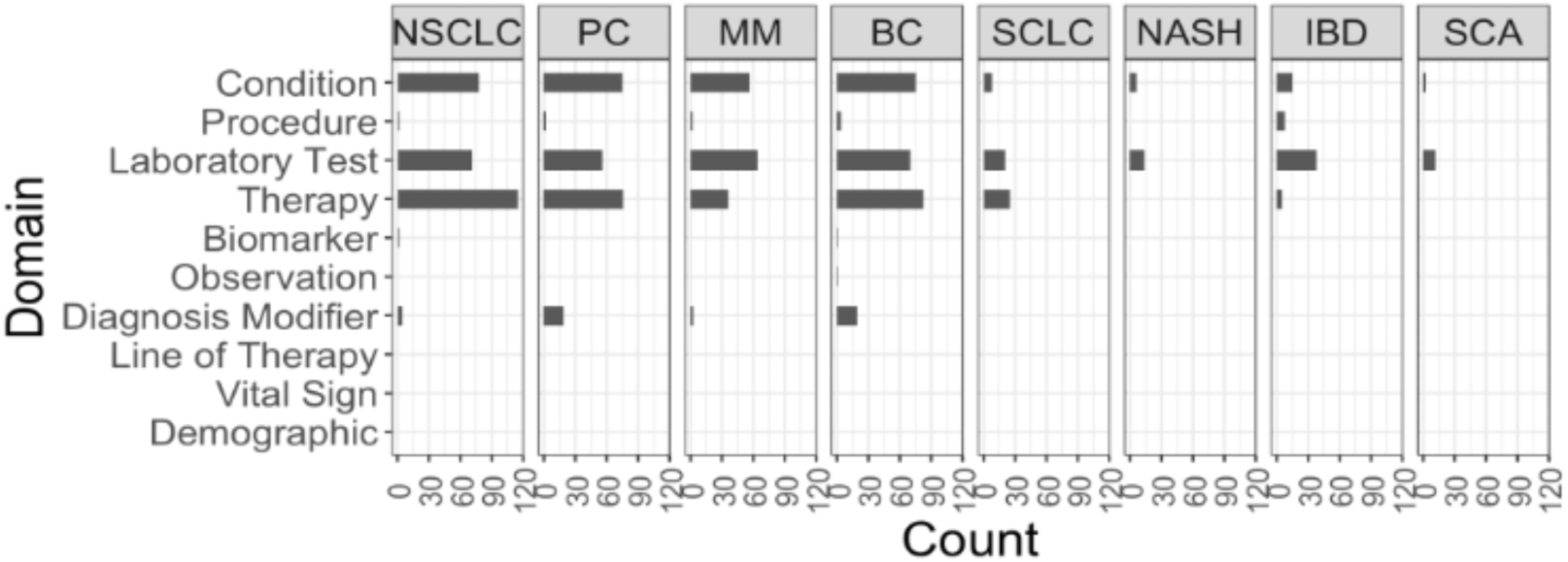
Clinical phenotypes in different clinical domains. (A) Distribution of annotated/normalized attributes across different clinical domains. (B) Distribution of annotated/normalized attributes of each disease across different clinical domains.

**Fig 3.**
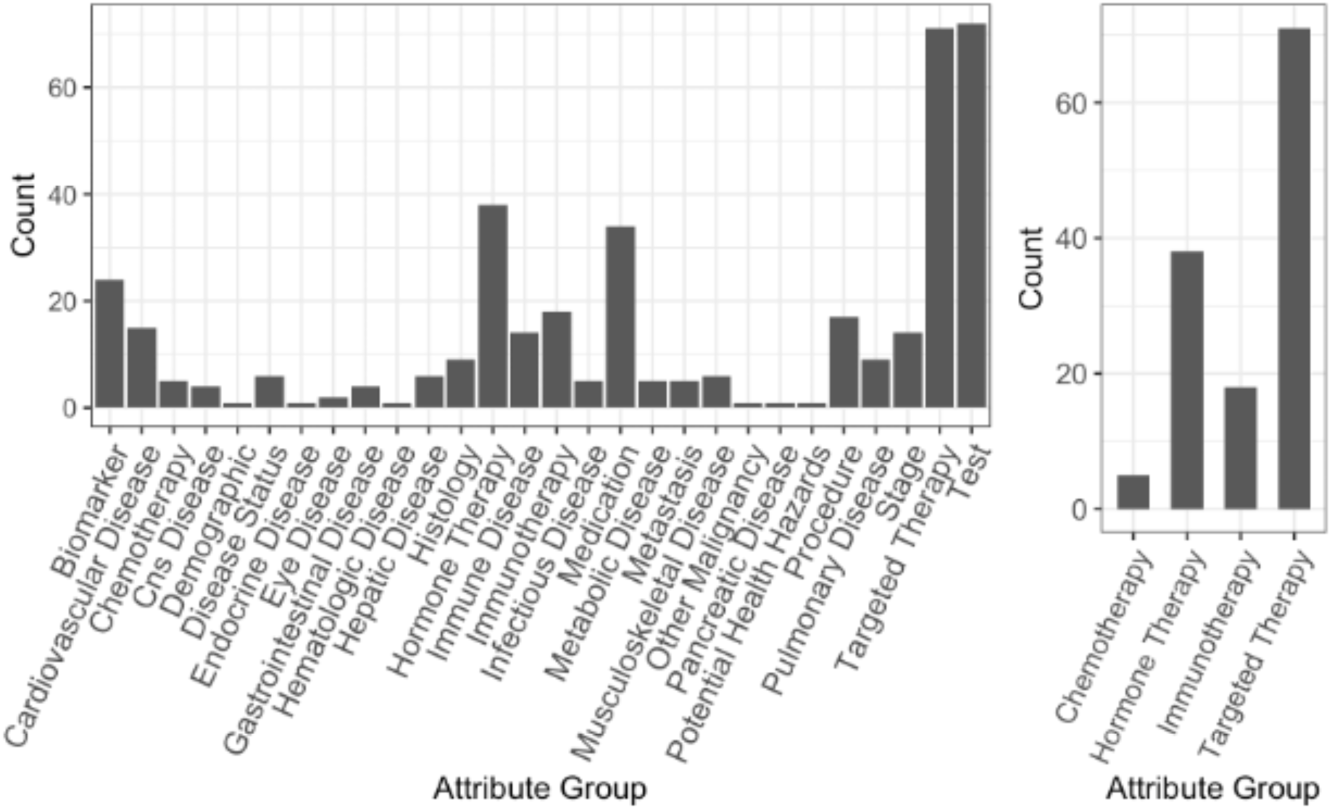
Clinical phenotypes in different attribute groups. (A) Distribution of annotated/normalized attributes across different attribute groups. (B) Distribution of annotated/normalized attributes across different modalities in clinical trials of cancer treatment.

Three of the top four EC attribute groups, medication, targeted therapy, and hormone therapy are related to the treatments: (i) treatments for comorbidities that are to be excluded, (ii) treatments that will interfere with the clinical trial, or (iii) treatments related to the diseases under study. The EC attribute groups within the therapy domain represent the cancer therapies that comprise regimen or medications used in cancer treatment. The EC attribute group, medication, includes drugs for treating cancer. The clinical trial attribute groups, targeted therapy and hormone therapy, are from the EC of cancer clinical trials. The drugs used in cancer treatment were regrouped into four attribute groups namely chemotherapy, targeted therapy, immunotherapy, and hormone therapy. Among these attribute groups, 51.91% (73/1231) belong to targeted therapy, 23.66% (31/131) belong to hormone therapy, 13.74% (18/131) belong to immunotherapy, and only 3.82% (5/131) belong to chemotherapy (Fig. 3B). The targeted therapy and hormone therapy are the most frequently mentioned treatment options for cancer.

We observed a set of commonly used attributes in the EC of clinical trials related to cancer (Fig. 4). These attributes describe the conditions (e.g., cardiovascular disease), treatments for these conditions (i.e. medication), previous line of therapy for cancer (e.g. chemotherapy), and lab test in the EC (blood, liver, and kidney function tests). These attributes may be considered when deciding on EC of cancer clinical trials.

**Fig. 4.**
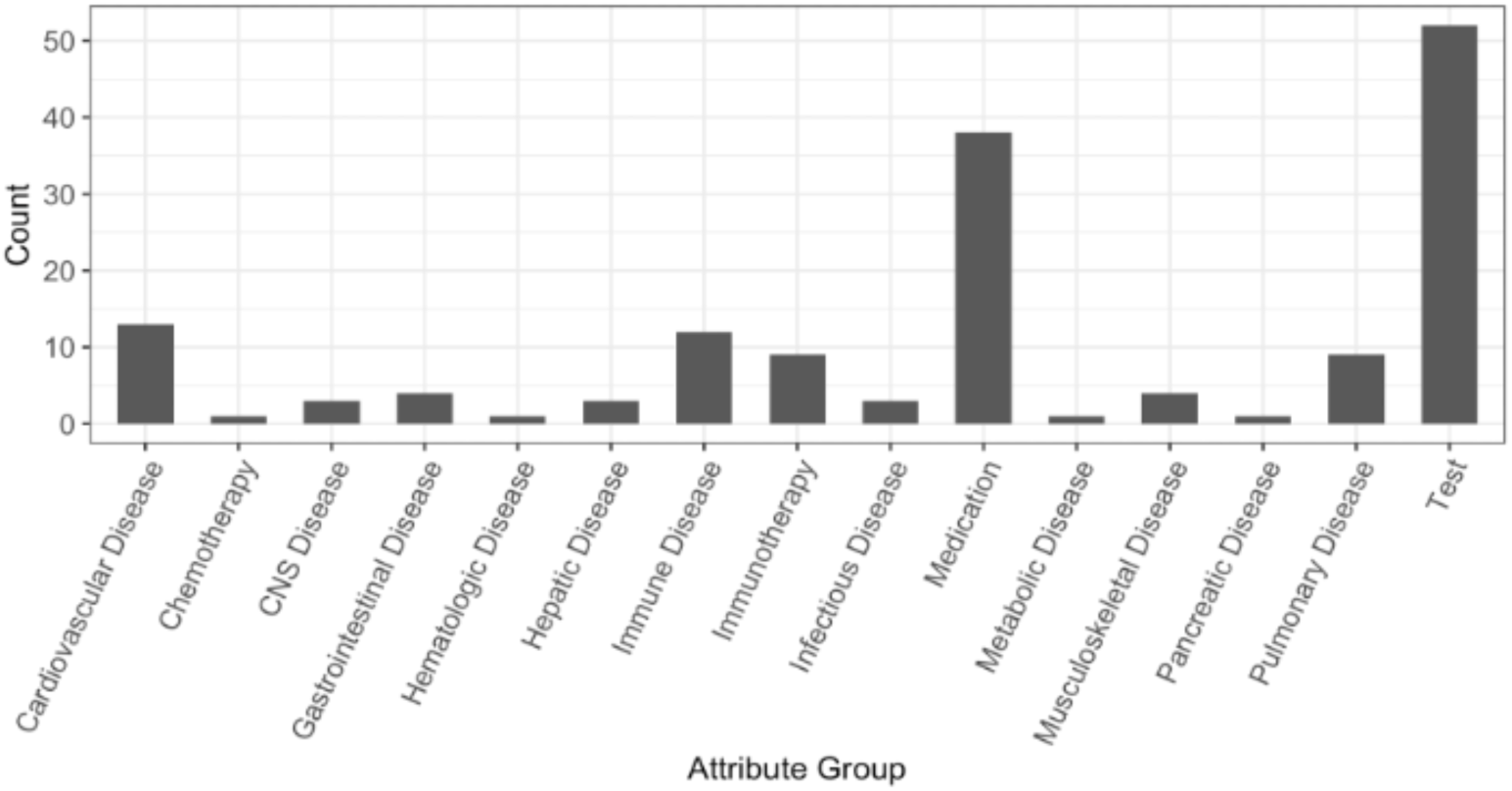
Common clinical phenotypes in clinical trials of cancer studies.

### Knowledge Base

Our knowledge base consists of two schema, clinical trials, and reference. Fig. 5 illustrates the schema and tables included in the knowledge base. The annotated and normalized attributes of EC of clinical trials from three domains, condition, procedure, and lab test were stored together in a master table under schema for clinical trial. The annotated and normalized clinical characteristics of EHR such as procedure, lab test, biomarker, and diagnosis modifier were stored in separate tables under schema for reference. The annotated and normalized clinical characteristics of EHR from two domains, therapy and observation were stored in one table. Each record can be queried using the attribute ID or attribute name.

**Fig. 5.**
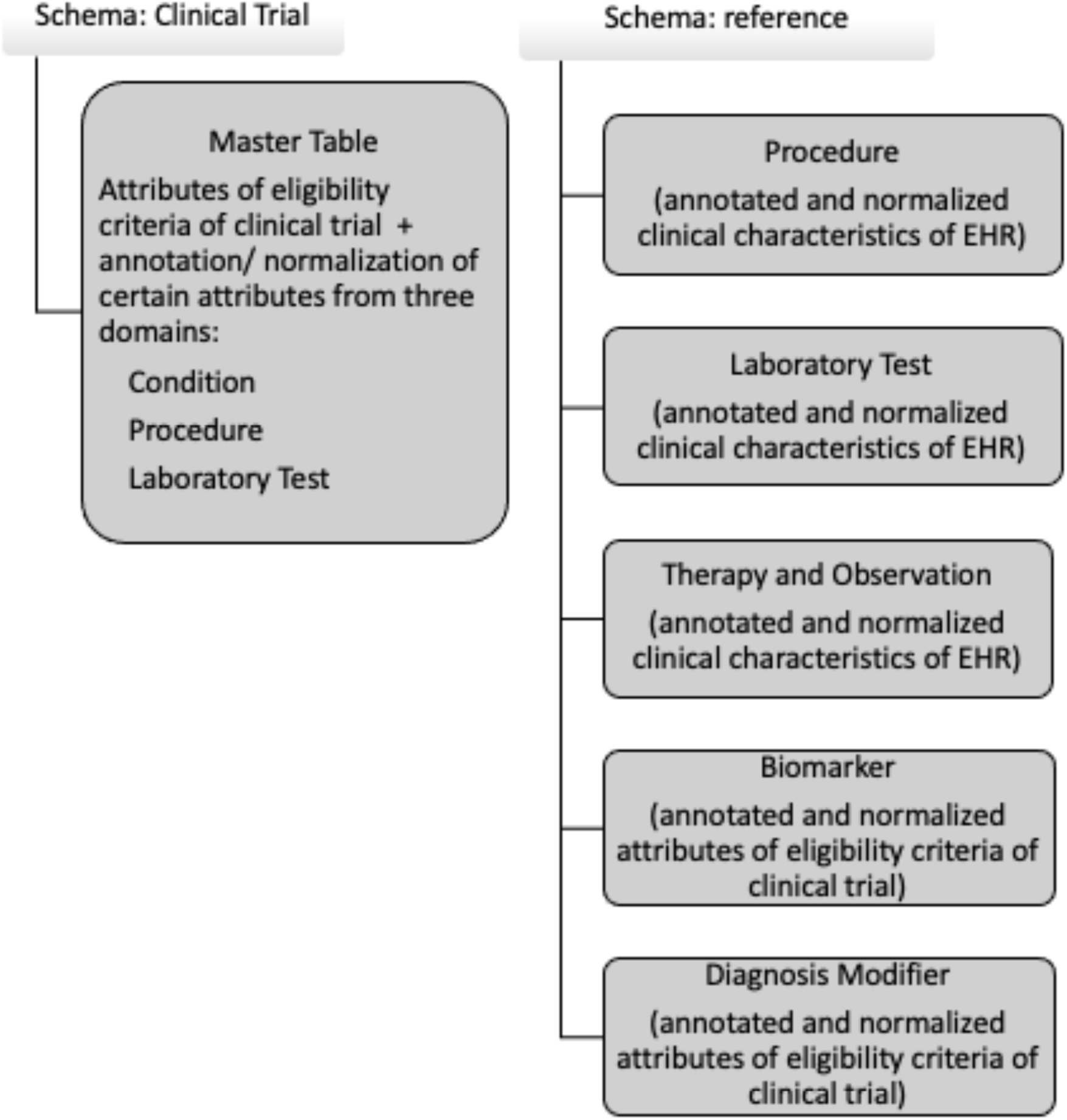
Knowledge base for annotated and normalized eligibility criteria attributes and normalized clinical characteristics of EHR.

#### Evaluation of semantic annotation and normalization

Table 3 shows the outcome of the quality control performed on a randomly selected subset of annotated and normalized clinical trial attributes of the EC of clinical trials and clinical characteristics of EHR within five domains, condition, procedure, lab test, therapy, and diagnosis modifier.

**Table 3.**
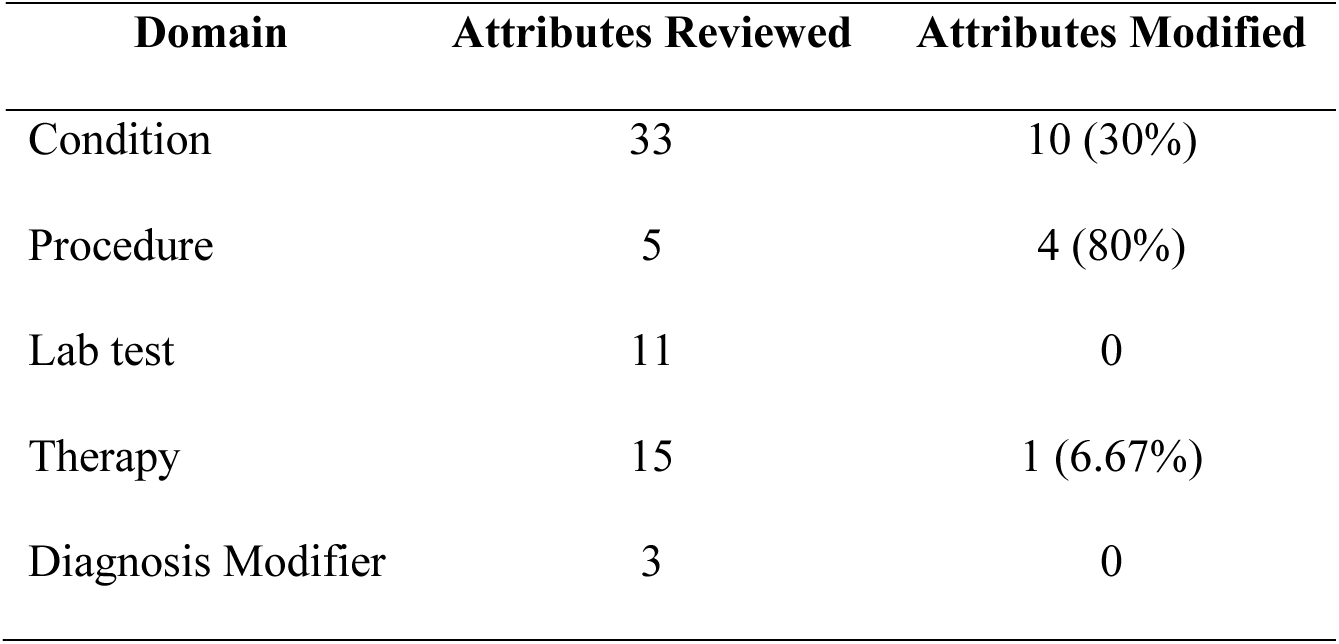
Quality control.

| Domain | Attributes Reviewed | Attributes Modified |
| --- | --- | --- |
| Condition | 33 | 10 (30%) |
| Procedure | 5 | 4 (80%) |
| Lab test | 11 | 0 |
| Therapy | 15 | 1 (6.67%) |
| Diagnosis Modifier | 3 | 0 |

#### Evaluation of Clinical Phenotyping Knowledge Base

The inter-rater agreement on the annotation of a random subset (89 out of 260 clinical trial attributes) of the knowledge base measured by Cohen’s Kappa coefficient is 0.82 (p = 0). The average performance score for patient matching measured by the F1-score among eight domains was 0.94, ranging from 0.82 to 1 (Table 4). The knowledge base was also successfully applied to EHR data from other institutes (data not shown) for patient pre-screening, suggesting its generalization capability.

**Table 4.**
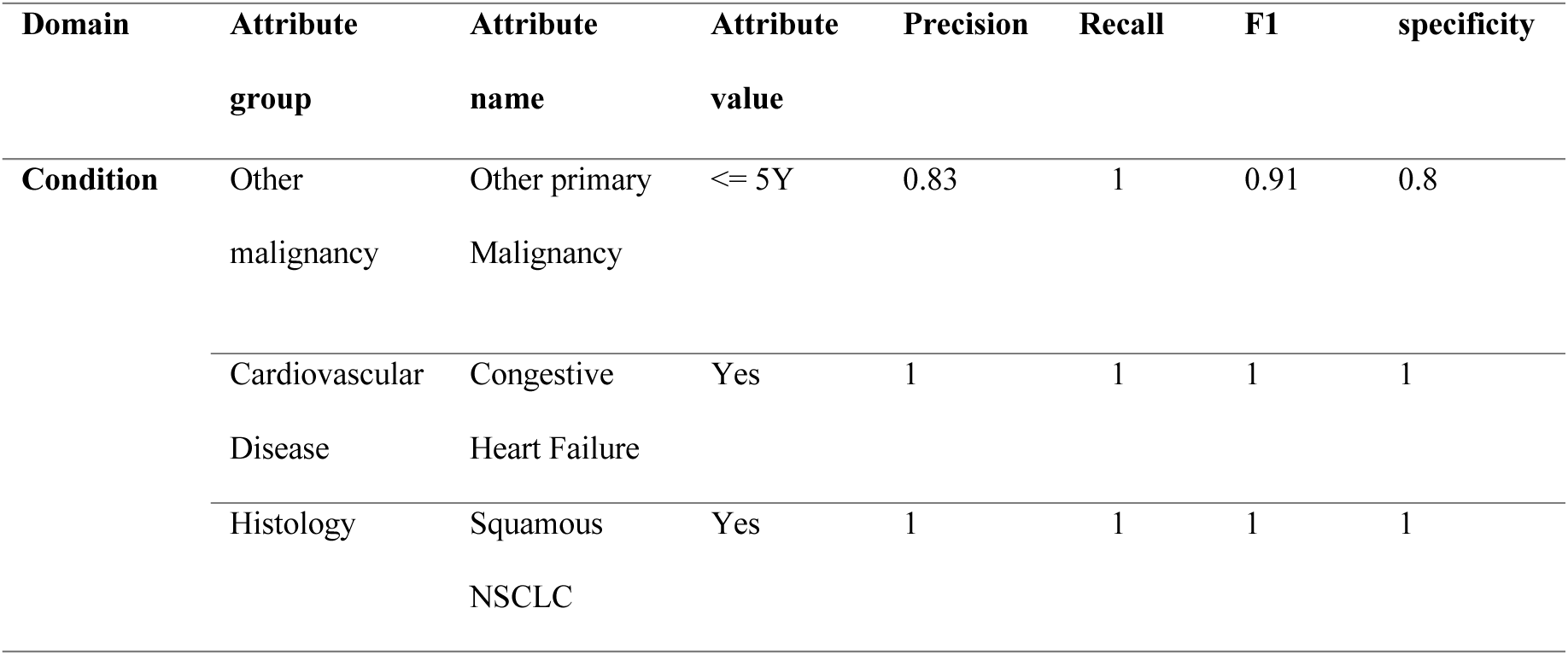
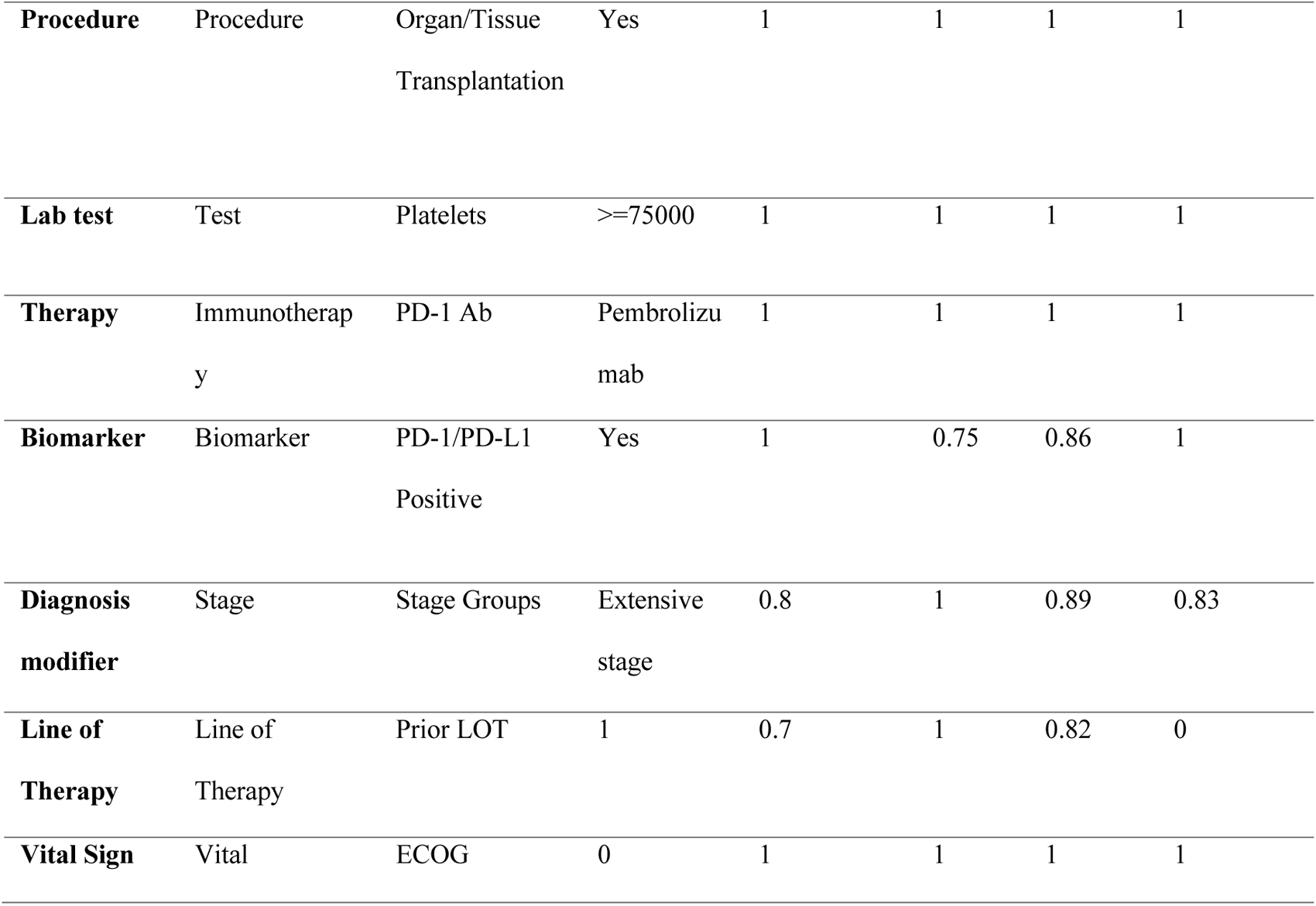
Evaluation of clinical attributes.

## Discussion

In this study, we built an intermediate representation of annotated and normalized attributes from the EC of clinical trials and the clinical characteristics found in EHR for clinical phenotyping. These annotated and normalized attributes facilitate the usability and interoperability of EHR data across multiple healthcare observational databases, making it easier to identify potentially eligible patients for clinical trials. The majority (87.74%) of the annotation and normalization work focused on three domains: condition, lab test, and therapy. These three domains were consistently mentioned in the EC of clinical trials across all the diseases analyzed. Therefore, the standardization of EHR data related to therapy, condition, and lab tests through standard terminology was prioritized to facilitate the development of an intermediate representation for EC clinical phenotyping.

In cancer clinical trials, targeted therapy and hormone therapy were more frequently mentioned than other types of therapy or modality. Immunotherapy had a smaller number of attributes compared to hormone therapy (47.37%) and targeted therapy (25.35%), but a greater number of attributes than chemotherapy (∼ 4%) (Figure 3). The last few decades have witnessed significant advancements in our understanding of molecular pathogenesis and the identification of novel disease-driven genetic disorders. These discoveries have led to the introduction of numerous targeted therapies, hormone therapy, and immunotherapy in cancer treatment. Currently, many of these therapies are being investigated in clinical trials and often aim to recruit subjects with relevant genetic alterations. Due to limited biomarker data in the current EHR database, a lower number of EC attributes from the biomarker domain was annotated and normalized (0.31 %) in this study. Expanding biomarker measurements in real-world would be beneficial for advancing precision medicine.

We phenotyped 92.37% of EC attributes (339 out of 367) in the domain of condition, procedure, lab test, and therapy. However, certain attributes including (i) CDAI (CD activity index), a diagnosis modifier attribute, (ii) fecal microbial transplantation, a procedure attribute, and (iii) NaPi2b targeted therapy, a therapy attribute, were not phenotyped due to unavailability of data in the structured EHR data in MSDW and VieCure. In future work, an alternative approach can be explored by leveraging data from the clinical notes for phenotyping. In our previous work (https://preprints.jmir.org/preprint/50800), we implemented advanced deep-learning NLP techniques using Conditional Random Fields (CRF) and Bi-directional Long Short-Term Memory to extract attributes from clinical trial EC. This pipeline can be further expanded to process clinical notes, enabling the automated phenotyping of attributes in clinical trial EC from huge text-based data.

### Limitation

Our study has several limitations. Firstly, limited biomarker data is available in the EHR database. Expanding biomarker measurements in real-world EHR data could improve the precision of phenotyping for clinical trials. Secondly, unavailability of certain eligibility criteria eligibility. Exploring alternative approaches, such as leveraging data from clinical notes, may help address this issue in future work. Thirdly, our normalization approach was carried out manually. The study acknowledges that using the billing code such as CPT for lab tests, and the standard encoding information such as the NDC (National Drug Code Dictionary) code for medications could automate the normalization process and accelerate the normalization of clinical characteristics. Additionally, leveraging the unique concept identifier (CUI) from UMLS Metathesaurus generated during data extraction using NLP can aid in automating the normalization of EC attributes. Moreover, our study focused on only one arm of clinical trials for clinical phenotyping. Future work aims to include attributes from every arm of the clinical trials to enhance the comprehensiveness of the analysis and further enrich the knowledge base.

## Conclusions

We developed a clinical trial phenotyping pipeline and knowledge base that maps clinical trial attributes to EHR clinical characteristics. This enables automated cohort selection for clinical trials and exhibits generalization across different institutes. Our approach complements standard terminologies, enhancing the normalization of clinical attributes and facilitating efficient patient matching for research.

## Data availability

The datasets generated and/or analyzed during the current study are not publicly available due to patient privacy, security, and the Health Insurance Portability and Accountability Act of 1996 (HIPAA) requirement but are available from the corresponding author on reasonable request.

## Authors’ contribution

K. Lee, Y. Mai, K. Raja, and X. Wang designed the study and wrote the manuscript. K. Lee and Y. Mai annotated clinical trial eligibility criteria, patient notes, and published knowledge bases. Y. Mai, Z. Liu, M. Ma, and T. Wang were involved in the data analysis. T. Jun, L. Ai, E. Calay, W. Oh, E. Schadt, X. Wang discussed the project and reviewed the manuscript.

